# Infoxmed2.0-27B: Instruction Tuning, Preference Alignment, and GRPO-Based Reward Model Training for Medical LLMs

**DOI:** 10.64898/2026.06.25.26356522

**Authors:** Jinlong Xie, Zhicai Guo, Hang Zhao, Hongjie Ni

**Affiliations:** School of Information Engineering, Sanming University, Sanming, China; Large Model Department, YiFu Technology Co., Ltd.; Medical Department, YiFu Technology Co., Ltd.

**Author notes:** Equal contribution (Jinlong Xie, Zhicai Guo).

**Keywords:** Medical Large Language Model, Instruction Fine-Tuning, Direct Preference Optimization, Group Relative Policy Optimization, Reward Model Training, LoRA

## Abstract

Large language models (LLMs) [1], [2] have demonstrated remarkable capabilities across general domains, yet their application in specialized medical contexts demands rigorous domain adaptation [3], [4]. We present Infoxmed2.0-27B, a medical foundation model built upon Qwen3.5-27B [5] through a comprehensive multi-stage post-training pipeline: (1) proprietary medical data synthesis from a MySQL database with MedicalCategoryTree organization, medical PhD team validation, Chinese RoBERTa [6] semantic deduplication, and API-assisted language refinement; (2) instruction supervised fine-tuning of Qwen3.5-27B via LoRA [7] (*r* = 8, *α* = 32) using MS-Swift [8], producing iterations Infoxmed2.0.0→ 2.0.2→ 2.0.4; (3) Direct Preference Optimization (DPO) [9] on 6,283 curated medical preference pairs [10] using DPO-RPO loss (*β* = 0.3, *α*_RPO_ = 0.1) across eight progressive training iterations (v0–v7); and (4) parallel Group Relative Policy Optimization (GRPO) [11]-based medical reward model training on Qwen3.5 combining internal rule-based reward functions with external DeepSeek signals. Comprehensive evaluations under a uniform LLM-as-Judge [12] framework with GPT-5.4 demonstrate 77.0% accuracy (mean quality score +7.18) on MedMCQA [10] and +2.59 on HLE, with pipeline progression from +6.69 (base) to +7.06 (SFT) to +7.18 (final).

## I. Introduction

The advent of large language models [2] built upon the Transformer architecture [1] has fundamentally transformed natural language processing, with healthcare emerging as one of the most impactful application domains [4], [13]. LLMs now demonstrate remarkable potential in clinical decision support, medical education, evidence-based medicine retrieval, and patient communication. However, general-purpose LLMs, despite their impressive breadth of knowledge, often lack the specialized medical knowledge, domain-specific reasoning patterns, and nuanced safety awareness required for high-stakes clinical scenarios [3]. This fundamental gap necessitates systematic domain adaptation that extends beyond simple prompt engineering or retrieval augmentation [14].

The research community has explored multiple post-training strategies for adapting LLMs to the medical domain. Super-vised fine-tuning (SFT) on medical instruction data [15], [16] establishes basic medical knowledge and instruction-following capabilities. Reinforcement Learning from Human Feedback (RLHF) [17], [18] leverages human preference judgments to align model outputs through reward model training and PPO-based policy optimization [19]. More recently, Direct Preference Optimization (DPO) [9] has emerged as a simpler and more stable alternative, directly optimizing the policy from preference pairs without requiring explicit reward model training or online sampling. Group Relative Policy Optimization (GRPO) [11] extends group-based optimization principles to reinforcement learning, enabling effective reward model training through group-relative comparisons within sampled response groups.

Despite these advances, most existing medical LLM efforts [16], [20] treat these techniques in isolation, lacking a cohesive framework that systematically orchestrates multiple training stages. Furthermore, training data quality remains a critical bottleneck, particularly in specialized domains where accuracy can have life-or-death implications [21], [22].

We introduce **Infoxmed2.0-27B**, a medical foundation model derived from Qwen3.5-27B [5] through a comprehensive multi-stage post-training pipeline. Our pipeline addresses the limitations of prior work through four synergistic components:

1. **High-quality medical data synthesis:** A proprietary pipeline extracting structured Q&A pairs from a MySQL database, organizing them through a hierarchical Medical-CategoryTree, enforcing quality through medical PhD team auditing and Chinese RoBERTa [6] semantic deduplication, and refining language through API-assisted optimization.
2. **Instruction supervised fine-tuning:** LoRA [7] finetuning (*r* = 8, *α* = 32, all-linear) of Qwen3.5-27B using the MS-Swift [8] framework, producing successive iterations from Infoxmed2.0.0 through Infoxmed2.0.4 with systematic evaluation at each stage.
3. **DPO-RPO preference alignment with progressive selection:** DPO [9] with RPO regularization on 6,283 curated medical preference pairs from MedMCQA [10] and HLE sources. Through eight progressive training runs (v0–v7), we track reward margins and select the checkpoint with optimal discriminative performance as the final model.
4. **GRPO-based reward model training:** In parallel with DPO, GRPO [11] trains a Qwen3.5-based medical reward model combining internal rule-based rewards with external DeepSeek signals, addressing the critical `num_labels=2` classification architecture challenge through dual-output reward computation.

We comprehensively evaluate Infoxmed2.0-27B on MedM-CQA [10] and HLE benchmarks using an LLM-as-Judge [12] framework with GPT-5.4. The model achieves 77.0% accuracy (mean quality score +7.18) on MedMCQA and +2.59 on HLE, with pipeline progression from +6.69 (base) to +7.06 (SFT) to +7.18 (final), comprehensively leading peer models including Baichuan-M2-32B, MedGemma-27B, Lingshu-32B, and HuatuoGPT-3-32B [20] under identical evaluation conditions.

## II. Related Work

### A. Medical Large Language Models

The adaptation of LLMs to the medical domain has attracted significant research attention [4], [13]. Med-PaLM 2 [13] achieved expert-level performance on medical question answering benchmarks through instruction prompt tuning on PaLM, demonstrating that fine-tuned general-purpose models can approach clinical expert quality. MedAlpaca [16] pioneered the open-source approach by leveraging GPT-3.5-generated [23] medical instruction data to fine-tune LLaMA models [24], making medical LLM capabilities more accessible. HuatuoGPT [20] further explored diverse medical data sources including clinical dialogues, examination questions, and medical literature to build a comprehensive Chinese-English bilingual medical model. On the Chinese biomedical NLP front, CBLUE [25] established a standardized evaluation benchmark covering multiple medical NLP tasks.

Despite these advances, existing medical LLMs face several persistent limitations. First, training data quality remains inconsistent—many approaches rely on LLM-generated synthetic data without rigorous professional validation, risking the propagation of factual errors and medical hallucinations [21]. Second, most works employ single-stage fine-tuning, missing the benefits of multi-stage optimization that systematically combines instruction tuning, preference alignment, and reward modeling. Third, the integration of reward models trained specifically for medical quality assessment is underexplored, with most systems relying solely on supervised objectives. Our work addresses these gaps through professionally validated data, progressive multi-stage training, and dedicated reward modeling.

### B. Parameter-Efficient Fine-Tuning

Low-Rank Adaptation (LoRA) [7] introduced a break-through approach by constraining weight updates to low-rank decompositions Δ*W* = *BA*, enabling adaptation with orders of magnitude fewer trainable parameters while maintaining performance comparable to full fine-tuning. QLoRA [26] further reduced memory requirements through 4-bit quantization, enabling fine-tuning of very large models on consumer hardware. In our work, we employ LoRA with rank *r* = 8 and *α* = 32, targeting all linear layers (all-linear configuration). This configuration provides an effective balance between adaptation capacity and parameter efficiency, requiring approximately 0.1% of the full parameter count for trainable weights while enabling thorough medical capability acquisition across both SFT and DPO stages.

### C. Preference Optimization and Reward Modeling

The challenge of aligning language model outputs with human preferences has driven significant methodological innovation. RLHF [17], [18] established the dominant paradigm: first training a reward model on human preference comparisons, then using PPO [19] to optimize the policy. DPO [9] fundamentally simplified this pipeline by reparameterizing the reward function in terms of the optimal policy, enabling direct optimization from preference pairs without explicit reward modeling. More recently, GRPO [11] introduced group-relative comparisons for reinforcement learning, enabling effective policy improvement through ranking-based optimization within response groups. This approach is particularly well-suited for training reward models, as it naturally captures the relative ordering of response quality. In our pipeline, DPO serves as the primary alignment mechanism for the main model, while GRPO is applied in parallel for reward model training, leveraging its group-relative comparison capability.

### D. Medical Benchmarks, Hallucination, and Infrastructure

MedMCQA [10] provides a large-scale multi-subject medical multiple-choice benchmark, while MMLU [27] includes medical subsets as part of broader knowledge assessment, and PubMedQA [28] targets biomedical reasoning with research-article-grounded questions. LLM-as-Judge [12], [29] enables scalable multi-dimensional quality assessment using strong LLMs like GPT-4 [23] as evaluators. Medical hallucination— the generation of plausible-sounding but factually incorrect medical information—poses particular risks in clinical contexts [21]. Constitutional AI [30] and retrieval augmentation [14] offer mitigation strategies. On the infrastructure side, DeepSpeed [31] provides ZeRO optimization stages, FlashAttention [32] reduces memory footprint through IO-aware computation, and vLLM [33] enables efficient inference serving through PagedAttention. These components collectively enable our pipeline to operate effectively on 4 ×A800 80GB GPUs.

## III. Methodology

### A. Overall Pipeline Architecture

Our post-training pipeline, illustrated in Fig. 1, comprises four sequential phases: (1) medical data synthesis under PhD team guidance from a proprietary MySQL database, (2) LoRA-based instruction SFT on Qwen3.5-27B, (3) parallel DPO preference alignment and GRPO reward model training, and (4) final model selection from the DPO stage based on optimal reward margin. The GRPO stage combines an internal reward function with an external DeepSeek reward model for auxiliary quality assessment.

**Fig. 1:**
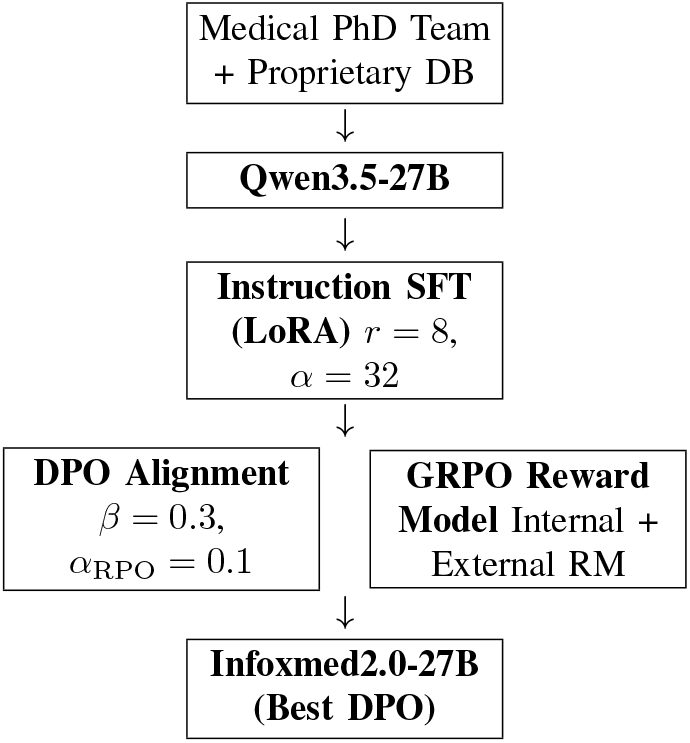
Post-training pipeline: Data synthesis → SFT → DPO ∥ GRPO-RM → best DPO checkpoint.

**Fig. 2:**
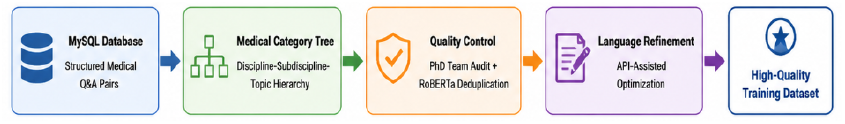
SFT medical instruction data synthesis pipeline: MySQL database extraction → Medical Category Tree organization → PhD team + RoBERTa quality control → API-assisted language refinement → high-quality training dataset.

**Fig. 3:**
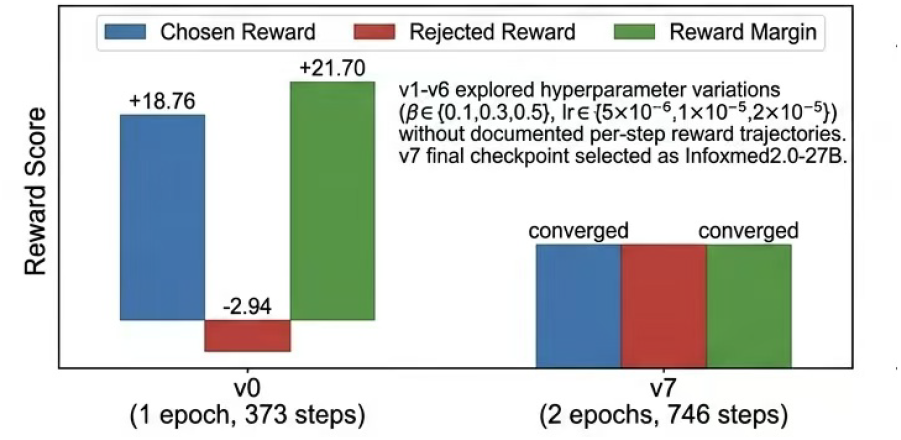
DPO training: v0 vs v7 reward comparison. v0 (1 epoch, 373 steps): chosen reward +18.76, rejected reward - 2.94, margin +21.70. v7 (2 epochs, 746 steps): reward margin converged and stable. v1–v6 explored hyperparameter variations (*β* ∈ { 0.1, 0.3, 0.5}, *lr* ∈ {5×10^−6^, 1×10^−5^, 2×10^−5^}) without documented per-step reward trajectories. The v7 final checkpoint was selected as Infoxmed2.0-27B.

**Fig. 4:**
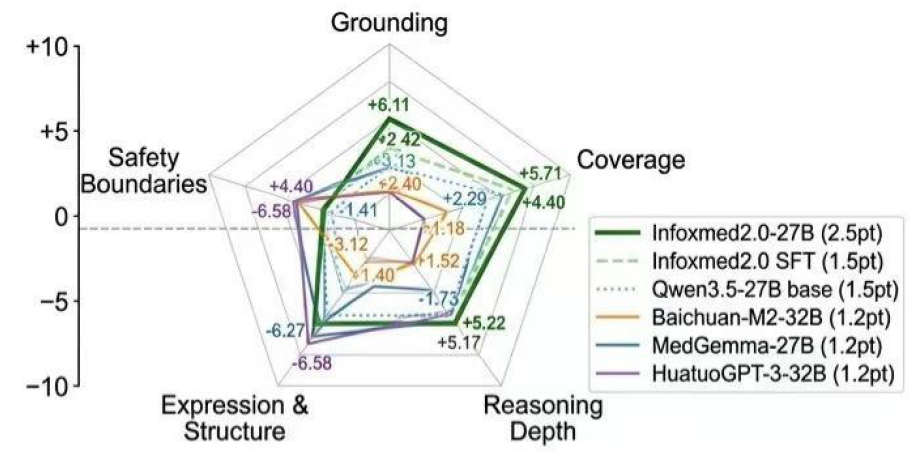
Radar chart comparison of six medical LLMs on the HLE benchmark across five evaluation dimensions. Infoxmed2.0-27B (dark green) leads in Expression & Structure (+6.11) and Safety Boundaries (+4.40), while all models exhibit negative Grounding scores, highlighting a common challenge in evidence consistency.

**Fig. 5:**
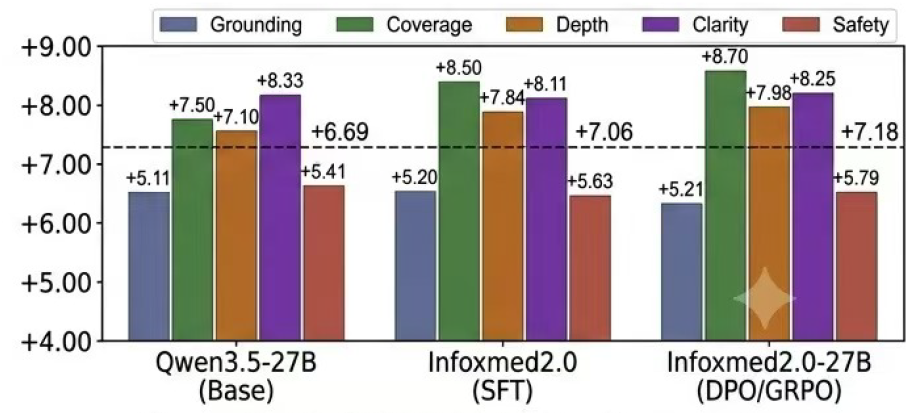
Pipeline progression on MedMCQA: quality scores across three training stages. The SFT stage substantially improves Coverage (+7.50 → +8.50) and Depth (+7.10 → +7.84). The DPO/GRPO stage further elevates the overall mean from +7.06 to +7.18 while recovering accuracy from 75% to 77%.

### B. Medical Instruction Data Synthesis

SFT training data quality is the single most critical determinant of downstream medical capability [15], [22]. Generic instruction datasets fail to capture the nuanced reasoning patterns, evidence-based methodology, and clinical safety considerations essential for medical AI systems. We therefore construct a comprehensive, multi-component data synthesis pipeline:

#### Data Sources and Extraction

Structured medical Q&A pairs are extracted from a proprietary MySQL database containing curated medical knowledge across multiple subdomains: anatomy, biochemistry, pharmacology, pathology, clinical diagnosis, and therapeutic protocols. Each record adheres to a standardized schema with question text, reference answers, supporting evidence, and domain classification tags. A dedicated extraction module queries the database with domain-specific filters and processes records through a thread-pool-based parallel pipeline with configurable batch sizes.

#### Medical Category Tree

A hierarchical MedicalCategoryTree structure organizes medical knowledge along three levels: broad disciplines (oncology, cardiology, neurology, endocrinology), subdisciplines (breast cancer, lung cancer, gastrointestinal oncology), and specific topics (molecular sub-types, treatment protocols, diagnostic biomarkers). This taxonomy ensures balanced coverage across medical subdomains, preventing the model from overfitting to specific disciplines.

#### Quality Assurance

A medical PhD team with clinical expertise conducts systematic quality audits covering: (i) medical accuracy of answers against authoritative references, (ii) logical completeness of reasoning pathways, and (iii) appropriate clinical boundary labeling. A Chinese RoBERTa model [6] performs semantic deduplication via pairwise cosine similarity scores, filtering samples above a threshold of 0.85 to ensure training set diversity. The parallel processing pipeline tracks quality statistics in real-time, capturing content length distributions, question type distributions, deduplication rates, and domain coverage metrics in a comprehensive statistics report (`stats_report.jsonl`).

#### Language Refinement

An API endpoint [34] calls a large language model to optimize the language organization of initial answers, improving naturalness and structural clarity while strictly preserving medical content accuracy. The refinement process operates with configurable batch sizes and timeout mechanisms for reliable processing at scale.

### C. Base Model: Qwen3.5-27B

We select Qwen3.5-27B [5], part of the Qwen family [35], for its strong multilingual capabilities across both English and Chinese, an extended 131K-token context window supporting long medical documents, and state-of-the-art reasoning performance on established benchmarks. The Transformer [1] architecture employs Rotary Position Embedding (RoPE), SwiGLU activation, Root Mean Square Layer Normalization (RMSNorm), and Grouped Query Attention (GQA) with 40 query heads and 8 key-value heads across 64 transformer layers. At 27 billion parameters, the model achieves an effective balance between representational capacity and computational requirements for deployment on our infrastructure of 4× NVIDIA A800 80GB GPUs.

### D. Instruction Supervised Fine-Tuning

We fine-tune Qwen3.5-27B using Low-Rank Adaptation (LoRA) [7], which constrains weight updates to low-rank decompositions Δ*W* = *BA* where *B* ∈ ℝ^*d*×*r*^ and *A* ∈ ℝ^*r*×*k*^ with rank *r* ≪min(*d, k*). We configure rank *r* = 8, scaling factor *α* = 32, and target all linear layers (all-linear), including query, key, value, and output projections in attention blocks as well as gate, up, and down projections in feed-forward networks.

Training employs the MS-Swift framework [8] with bfloat16 mixed precision for numerical stability. We use the AdamW fused optimizer with learning rate 1 × 10^−5^, cosine learning rate schedule with 10% warmup steps, and weight decay of 0.1 for regularization. Distributed training runs on 4× NVIDIA A800 80GB GPUs using DeepSpeed ZeRO-2 [31], achieving an effective batch size of 16 through per-device batch of 2 with gradient accumulation over 2 steps. Training proceeds for 2 epochs with a maximum sequence length of 3,072 tokens. Model checkpoints are saved every 50 steps (save total limit 5) with evaluation on a 5% held-out validation split. A self-hosted WandB instance provides real-time experiment tracking.

Through multiple SFT rounds with iterative data quality improvements and hyperparameter adjustments, we produce successive model versions: Infoxmed2.0.0 (initial baseline), Infoxmed2.0.2 (incorporating expanded training data and refined hyperparameters), and Infoxmed2.0.4 (the final SFT model serving as foundation for subsequent DPO and GRPO stages). Each iteration undergoes systematic evaluation to quantify improvements.

### E. Direct Preference Optimization with RPO Regularization

The DPO stage [9] aligns model outputs with medical quality preferences through direct optimization from preference pairs, eliminating the need for explicit reward model training and online sampling.

#### Preference Data Construction

We construct 6,283 medical preference pairs (`dpo_medical_train_v2.jsonl`) from MedMCQA [10] and HLE question sources. Each pair contains a high-quality *chosen* response (full reasoning chains, structured output with clear sectioning, explicit evidence citations, and medically accurate content) and a lower-quality *rejected* response exhibiting one of three error types [21]: Vague (insufficient detail, avoidance of definitive answers, superficial analysis), Hallucination (fabricated medical facts, imaginary clinical trials, plausible-sounding but factually unsupported claims), and Incomplete or Wrong (truncated responses, missing critical diagnostic information, incorrect answer options). Each pair includes comprehensive metadata with source dataset, error type classification, and five-dimensional quality scores. A data generation toolchain (`generate_dpo.py`, `filter_dpo.py`, `analyze_dpo_dataset.py`) automates construction, filtering, and statistical analysis.

#### Training Configuration

MS-Swift [8] DPO implementation with LoRA (*r* = 8, *α* = 32, all-linear). DPO-specific hyperparameters: *β* = 0.3 (controls preference strength by modulating the logistic function’s sensitivity to policy ratio differences) and *α*_RPO_ = 0.1 (RPO regularization coefficient that adds SFT loss on chosen responses to anchor the model and prevent excessive deviation from the SFT-quality baseline). Training is distributed across 4× A800 80GB GPUs with DeepSpeed ZeRO-2 [31], effective batch size 16, learning rate 1 × 10^−5^, and 2 training epochs. Maximum sequence length is 3,072 tokens with left truncation strategy (`truncation_strategy=left`) to preserve the tail portions of responses. Dataset preprocessing employs 4 parallel workers with caching enabled. A 5% validation split (314 samples) monitors generalization, with evaluation and checkpoint saving every 50 steps.

The DPO-RPO loss combines preference optimization with SFT regularization [9]:

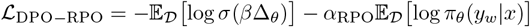

where 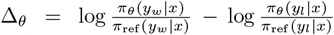 quantifies the policy’s relative preference for chosen over rejected responses.

#### Progressive Training and Model Selection

DPO training proceeds through 8 iterative runs (v0 through v7). The initial v0 run (1 epoch, 373 steps, per-device batch 1, gradient accumulation 8) achieves a strong reward margin of 21.70 (chosen 18.76, rejected −2.94) with training loss converging to approximately 0.0325. Subsequent versions systematically explore hyperparameter variations. Version v7 (2 epochs, 746 steps) represents the final configuration. The checkpoint with optimal discriminative performance—maximum chosen– rejected margin without rejected reward inflation—is selected as the final model. A full-parameter DPO variant using Deep-Speed ZeRO-3 [31] with `padding_free` optimization and FlashAttention [32] reduces per-GPU memory from 57GB to 47GB and accelerates training from 3.32 to 1.90 seconds per iteration.

### F. GRPO-Based Medical Reward Model Training

In parallel with DPO, we apply Group Relative Policy Optimization (GRPO) [11] to train a dedicated medical reward model based on Qwen3.5 [5] with LoRA [7]. The reward signal adopts a combined architecture of internal reward functions and external reward model.

#### Internal Reward Function

Dense granular rule-based rewards encode medical quality along multiple dimensions with hierarchical granularity: field-level correctness (points for selecting appropriate search fields such as Title/Abstract), boolean operator accuracy (points for proper use of AND, OR, and NOT operators), syntax validity (points for correct query structure and bracket matching), and retrieval success (points for whether information-seeking actions produce meaningful results). A critical design choice is the explicit avoidance of reward clipping. Dense, continuous reward signals preserve the gradient of improvement, enabling the model to learn sophisticated quality discrimination rather than converging to locally optimal but globally suboptimal behaviors [19].

#### External Reward Model

DeepSeek provides holistic quality assessments on a 0–1 scale, evaluating overall medical accuracy, coherence, clinical appropriateness, and evidence integration. The combination of internal granular rules (providing specific, interpretable feedback on atomic quality components) and external holistic assessment (capturing global quality patterns) addresses the fundamental limitation of single-mechanism reward approaches.

#### num_labels=2 **Architecture Challenge**

A significant technical challenge emerged from Qwen3.5’s classification model architecture. Despite explicitly setting `num_labels=1` during model loading via the transformers library [36], the base model’s built-in configuration (`num_labels: 2, id2label: {0: ‘LABEL_0’, 1: ‘LABEL_1’}`) takes precedence, producing a [2, 4096] output tensor rather than the expected [1, 4096]. Our adaptation strategy works with rather than against this constraint: the `compute_loss` function extracts positive score *s*_pos_ and negative score *s*_neg_ from dual outputs, computing *r* = *s*_pos_ −*s*_neg_ with pairwise training loss ℒ = − log *σ*(*r*_chosen_− *r*_rejected_). LoRA employs `modules_to_save=[“score”]` for correct score layer preservation during adapter saving.

#### Training Configuration and Results

Single-GPU training with the VERL [37] framework (`algorithm.adv_estimator=grpo`). Hyperparameters: rollout sample count *n* = 4, KL penalty *λ*_KL_ = 0, entropy coefficient = 0, PPO clipping range [0.2, 0.3], learning rate 1 ×10^−6^, 2 epochs, max prompt 8,192 tokens, max response 2,048 tokens. Rollouts use vLLM [33] (GPU memory utilization 0.6) with FSDP parameter and optimizer offloading. The trained reward model achieves 46% pairwise accuracy (mean chosen reward 2.552, mean rejected reward 2.627, gap − 0.075), demonstrating basic but functional discriminative capability.

## IV. Experimental Setup

### A. Training Infrastructure

4× NVIDIA A800 80GB GPUs, Docker (Ubuntu 22.04, CUDA 12.6.3, PyTorch 2.7.1, vLLM 0.10.1.1 [33], MS-Swift 3.8.1 [8]). GRPO: VERL 0.5.0 [37]. SFT data: MySQL, Dify [34] API. Self-hosted WandB for tracking.

### B. Evaluation Benchmarks

**MedMCQA [10]:** Multi-subject medical MCQ (Anatomy, Biochemistry, Pharmacology, Pathology), 200 questions. **HLE (Hard Logic Evaluation):** Deep medical reasoning, 200 questions (multimodal open-ended; accuracy not reported).

### C. Evaluation Methodology

LLM-as-Judge [12] (GPT-5.4), five dimensions (−10 to +10): Grounding (Evidence Consistency), Coverage, Reasoning Chain Completeness, Expression & Structure, Safety Boundaries. MedMCQA additionally reports accuracy via exact match.

## Results and Analysis

### A. MedMCQA Evaluation

Table I shows 77% accuracy, Coverage +8.70, Expression & Structure +8.25.

**TABLE I:** MedMCQA results (200 questions).

| Metric | Score |
| --- | --- |
| Accuracy | 77.00% |
| *Five-Dimension Scores* |  |
| Grounding | +5.21 |
| Coverage | +8.70 |
| Reasoning Chain Completeness | +7.98 |
| Expression & Structure | +8.25 |
| Safety Boundaries | +5.79 |
| Overall Mean | +7.18 |

### B. HLE Evaluation

Table II: 95.5% accuracy, Grounding − 3.08 (fabrication on out-of-domain questions [21]), Expression & Structure +6.11.

**TABLE II:** HLE results (200 questions).

| Metric | Score |
| --- | --- |
| Accuracy | 95.50% |
| *Five-Dimension Scores* |  |
| Grounding | −3.08 |
| Coverage | +3.10 |
| Reasoning Chain Completeness | +2.42 |
| Expression & Structure | +6.11 |
| Safety Boundaries | +4.40 |
| Overall Mean | +2.59 |

### C. Comprehensive Peer Model Comparison

Identical conditions [12]: Baichuan-M2-32B, MedGemma-27B, Lingshu-32B, HuatuoGPT-3-32B [20], Qwen3.5-27B base, Infoxmed2.0 SFT. Table III: Infoxmed2.0-27B leads +7.18, 77.0% accuracy. Pipeline: + 6.69 → +7.06 → +7.18. Table IV: HLE comparison (accuracy not reported). Infoxmed2.0-27B +2.59, Expression & Structure +6.11, Safety +4.40 (first). All models negative Grounding; Infoxmed2.0-27B (−3.08) slightly worse than base (−1.41).

**TABLE III:**
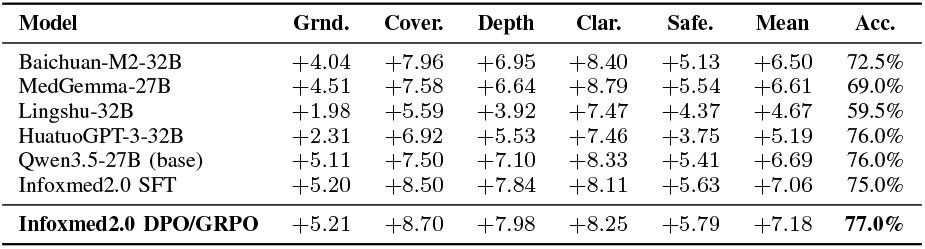
Peer comparison on MedMCQA (200 questions). Bold: our models.

**TABLE IV:** Peer comparison on HLE (200 questions). Bold: our models.

| Model | Grnd. | Cover. | Depth | Clar. | Safe. | Mean |
| --- | --- | --- | --- | --- | --- | --- |
| Baichuan-M2-32B | -3.12 | +1.40 | +1.18 | +2.92 | +1.52 | +0.78 |
| MedGemma-27B | -6.27 | +2.00 | -1.73 | +5.22 | +3.10 | +0.47 |
| Lingshu-32B | -6.65 | +0.38 | -3.90 | +3.96 | +3.02 | -0.64 |
| HuatuoGPT-3-32B | -6.58 | +2.40 | -0.09 | +5.17 | +2.27 | +0.64 |
| Qwen3.5-27B (base) | -1.41 | +2.13 | +2.29 | +4.40 | +3.21 | +2.12 |
| Infomed2.0 SFT | -2.48 | +3.13 | +2.50 | +5.71 | +4.13 | +2.60 |
| **Infomed2.0 DPO/GRPO** | **-3.08** | **+3.10** | **+2.42** | **+6.11** | **+4.40** | **+2.59** |

### D. Reward Model and GRPO Observations

Reward model: 46% pairwise accuracy [11]. Internal– external combined strategy outperforms single mechanisms. Dense rewards critical; PPO clipping sensitivity; ∼ 56GB minimum VRAM.

## VI. Discussion

SFT [7], [15] with professional data synthesis (Coverage +8.70, Expression +8.25). DPO [9] progressive checkpoint selection. GRPO [11] combined rewards with num labels=2 adaptation [36]. Limitations: (1) HLE Grounding −3.08 vs base −1.41 [21]; (2) 46% reward accuracy [18]; (3) Imperfect safety [30]; (4) 6,283 DPO pairs [9]; (5) Two benchmarks [10],[27].

## VII. Conclusion

Infoxmed2.0-27B: medical PhD-guided data synthesis, LoRA SFT [7], DPO-RPO [9] (v0–v7), GRPO reward model [11] with combined internal–external signals and num labels=2 adaptation [36]. Best DPO checkpoint as final model. 77.0% accuracy (+7.18) MedMCQA, +2.59 HLE, leading peers [12].

## Data Availability

https://www.modelscope.cn/datasets/InfoxmedModel/Infoxmed2.0_data

## Acknowledgements

We thank the Qwen team [5], DeepSpeed [31], MS-Swift [8], VERL [37] communities.

